# The reduction of sleep-like perilesional cortical dynamics underlies clinical recovery in stroke

**DOI:** 10.1101/2024.03.16.24304272

**Authors:** S. Sarasso, S. D’Ambrosio, S. Russo, L. Bernardelli, G. Hassan, A. Comanducci, P. De Giampaulis, L. Dalla Vecchia, J. Lanzone, M. Massimini

**Author notes:** These authors equally contributed (S. Sarasso, S. D’Ambrosio). **Correspondence should be sent to:**.

## Abstract

**Introduction:** Recent studies have shown that, following brain injury, sleep-like cortical dynamics intrude into wakefulness, potentially contributing to brain network disruption and behavioral deficits.

**Aim:** We employ TMS in combination with EEG to detect these dynamics and assess their impact on brain networks and clinical evolution in awake stroke patients.

**Methods:** Twelve patients with subacute unilateral ischemic cortical stroke underwent a longitudinal study with two assessments (t0 and t1), including clinical evaluation using the National Institutes of Health Stroke Scale (NIHSS) and TMS-EEG recordings targeting perilesional and contralesional cortical sites. Parameters such as slow wave amplitude (SWa), high-frequency power (HFp) suppression, and the Perturbational Complexity Index-state transition (PCIst) were analyzed to quantify sleep-like cortical dynamics and their network-level consequences.

**Results:** Results demonstrated a significant clinical improvement (NIHSS score: 7.16±0.73 at t0, 4.33±0.74 at t1; W=78, P<0.001). Perilesional SWa and HFp suppression decreased significantly at t1 compared to t0 (T(11)=3.05, P=0.01 and T(11)=-3.39, P<0.01, respectively), along with recovery of PCIst values (T(11)=-2.35, P=0.04). Importantly, both the dissipation of sleep-like perilesional cortical dynamics and the recovery of network-level interactions correlated with patients’ clinical improvement (Spearman ρ=0.62, P=0.03; ρ=-0.68, P=0.01, respectively).

**Conclusion:** These findings underscore the potential of TMS-EEG as an objective measure of neurological evolution and suggest targeting sleep-like cortical dynamics as a viable strategy for post-stroke neuromodulation and rehabilitation.

## Introduction

The consequences of focal brain lesions are known to extend beyond the site of structural injury [1]. The presence of altered neuronal activity outside the area of structural injury was postulated already more than 100 years ago, and termed diaschisis by Costantin Von Monakow [2]. Subsequent experimental work, substantiated diaschisis as the depression of perfusion/metabolic function [3] involving structurally intact brain areas connected to the injury [4], and showed its impact on behavioral functions [5]. Notwithstanding more recent multimodal imaging assessments [6–9] confirming and complementing these observations, the electrophysiological determinants of diaschisis are still elusive.

From the electrophysiological standpoint, the presence of EEG slowing in cortical areas surrounding brain lesions has also been known for almost a century [10], albeit never connected to the concept of functional network disruption after brain injury and diaschisis. Recent works involving Transcranial Magnetic Stimulation coupled with EEG (TMS-EEG) and intracranial recordings confirmed that slow waves can intrude into wakefulness following brain injuries and added important elements. First, they showed that post-lesional slow waves are indistinguishable from sleep slow waves, being also underpinned by cortical bistability [11], i.e. the tendency of cortical neurons to fall in a silent period (OFF-period) after an initial activation. Second, they demonstrated that sleep-like bistability is more prominent in areas that are near, or connected to, the lesion [12]. Third, they showed that post-lesional bistability and OFF-periods can impair neuronal and network interactions [11, 13], with a potentially significant impact on behavior [14].

Despite its potential explanatory power and clinical impact, the relationship between perilesional slow waves, the associated disruption of cortical interactions and their effects on behavior has never been systematically assessed. Here, we fill this gap by performing longitudinal assessments in 12 subacute patients affected by unilateral ischemic cortical stroke in whom we measured the EEG response to TMS in addition to spontaneous EEG-derived quantitative metrics. We investigate the presence and the evolution of perilesional EEG markers of cortical bistability (i.e. the presence of TMS-evoked perilesional EEG slow waves associated with OFF-periods) and measure their impact on local cortical interactions. Crucially, we assess whether the evolution of such measures was paralleled by the clinical evolution of the patients as measured by validated quantitative scales.

We show that the reduction of perilesional cortical bistability over time is proportional to the clinical improvement and is paralleled by a recovery of cortical interactions, thus suggesting an association between the functional state of the cortical tissue surrounding the lesion and behavioral functions. Importantly, such clinically relevant association could be readily revealed by probing the cortex with TMS-EEG but could not be systematically detected by spontaneous quantitative EEG measures.

The present results suggest that assessing the intrusion of sleep-like cortical dynamics by a perturbational perspective and tracking its evolution over time can provide an objective read-out of the patients’ neurological evolution. Further, they open the perspective that reducing cortical bistability during the subacute phase post-injury may represent a tractable neurophysiologically-grounded target for treatment and rehabilitation.

## Materials and methods

### Participants and Experimental procedures

We recruited a total of 12 participants (5 females, median=71.5y, IQR=12.5y) affected by unilateral ischemic stroke of different lesion volume (mean ± SEM: 53.76 ± 11.73cm^3^) and location (see Table 1). All patients underwent two experimental sessions, one at the beginning (t0) and one at the discharge of the rehabilitation stay (t1) (mean duration ± SEM: 85.92 ± 25.12 days). During each experimental session, patients were clinically evaluated using the National Institutes of Health Stroke Scale (NIHSS; [15]). In addition, two TMS-EEG measurements were performed with TMS targeted to intact cortical portions of both the affected (perilesional stimulation site) and the unaffected (contralesional stimulation site) hemispheres. CT scans were acquired at t0 to operationalize the selection of TMS targets [13] and to guide neuronavigation during both experimental sessions. Specifically, the perilesional stimulation site involved a portion of the same Brodmann Area (BA; see Table 1) including the anatomical lesion within 2 cm of its boundaries, while the contralesional stimulation site corresponded to the homologue contralateral cortical area. When stimulating the primary motor cortex (BA4) we explicitly avoided inducing visible muscle twitches (hand/arm, face, shoulder/ trunk and foot/leg) to avoid TMS-evoked EEG potentials (TEPs) contamination by the sensory re-entry of proprioceptive feedback [16].

**Table 1.**
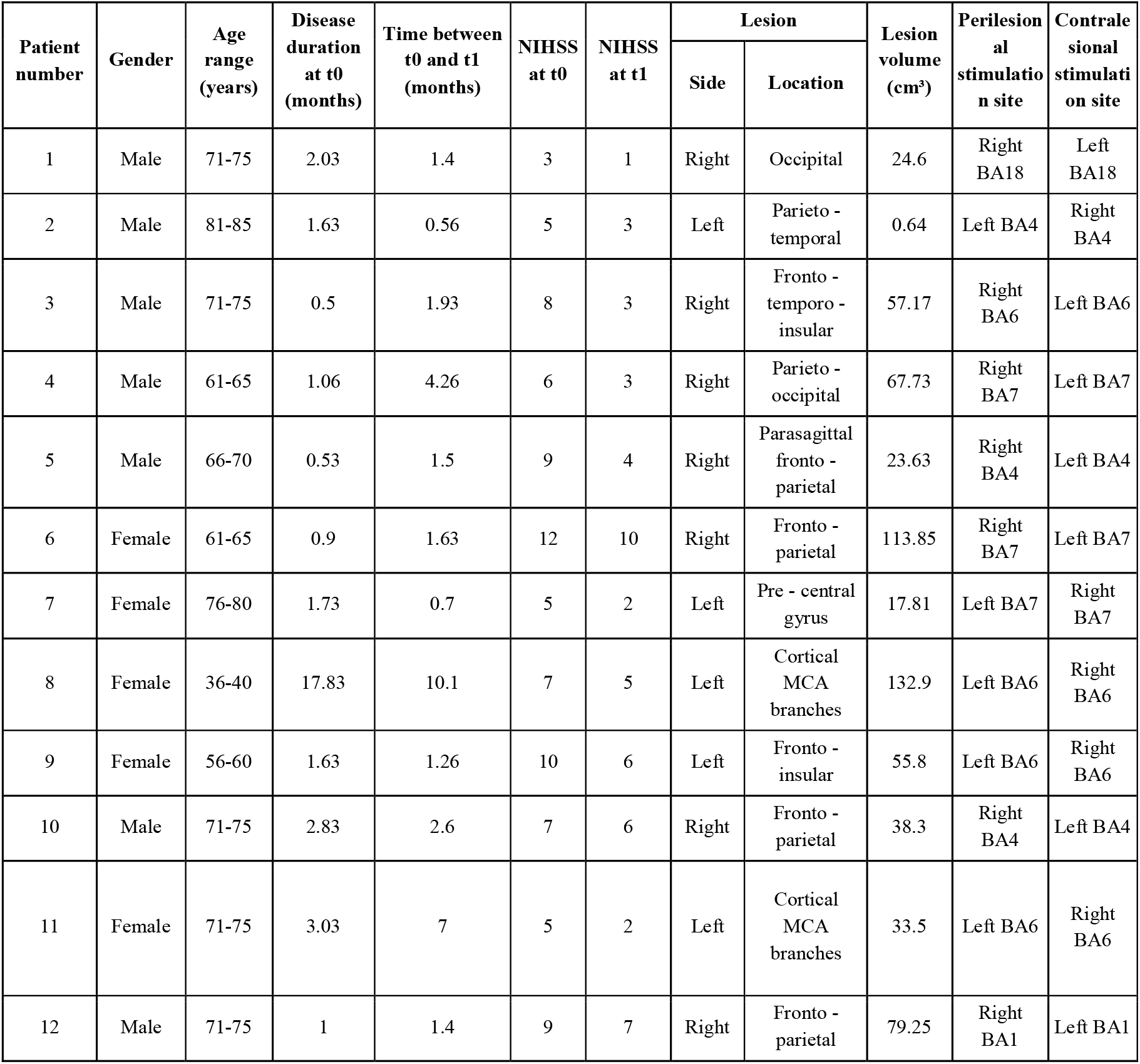
Patients’ demographic and clinical data. NIHSS: NIH Stroke Scale. BA: Brodmann Area

| Patient number | Gender | Age range (years) | Disease duration at t0 (months) | Time between t0 and t1 (months) | NIHSS at t0 | NIHSS at t1 | Lesion |  | Lesion volume (cm<sup>3</sup>) | Perilesional stimulation site | Contralateral stimulation site |
| --- | --- | --- | --- | --- | --- | --- | --- | --- | --- | --- | --- |
|  |  |  |  |  |  |  | Side | Location |  |  |  |
| 1 | Male | 71-75 | 2.03 | 1.4 | 3 | 1 | Right | Occipital | 24.6 | Right BA18 | Left BA18 |
| 2 | Male | 81-85 | 1.63 | 0.56 | 5 | 3 | Left | Parieto - temporal | 0.64 | Left BA4 | Right BA4 |
| 3 | Male | 71-75 | 0.5 | 1.93 | 8 | 3 | Right | Fronto - temporo - insular | 57.17 | Right BA6 | Left BA6 |
| 4 | Male | 61-65 | 1.06 | 4.26 | 6 | 3 | Right | Parieto - occipital | 67.73 | Right BA7 | Left BA7 |
| 5 | Male | 66-70 | 0.53 | 1.5 | 9 | 4 | Right | Parasagittal fronto - parietal | 23.63 | Right BA4 | Left BA4 |
| 6 | Female | 61-65 | 0.9 | 1.63 | 12 | 10 | Right | Fronto - parietal | 113.85 | Right BA7 | Left BA7 |
| 7 | Female | 76-80 | 1.73 | 0.7 | 5 | 2 | Left | Pre - central gyrus | 17.81 | Left BA7 | Right BA7 |
| 8 | Female | 36-40 | 17.83 | 10.1 | 7 | 5 | Left | Cortical MCA branches | 132.9 | Left BA6 | Right BA6 |
| 9 | Female | 56-60 | 1.63 | 1.26 | 10 | 6 | Left | Fronto - insular | 55.8 | Left BA6 | Right BA6 |
| 10 | Male | 71-75 | 2.83 | 2.6 | 7 | 6 | Right | Fronto - parietal | 38.3 | Right BA4 | Left BA4 |
| 11 | Female | 71-75 | 3.03 | 7 | 5 | 2 | Left | Cortical MCA branches | 33.5 | Left BA6 | Right BA6 |
| 12 | Male | 71-75 | 1 | 1.4 | 9 | 7 | Right | Fronto - parietal | 79.25 | Right BA1 | Left BA1 |

During all measurements, patients were awake sitting comfortably and with their eyes open. Vigilance was monitored using EEG to promptly detect any signs of drowsiness. The study conformed to the standards set by the Declaration of Helsinki; ethical approval was obtained from the ethical committee of IRCCS Istituti Clinici Scientifici Maugeri, Pavia, Italy (# 2286CE). Each patient gave written informed consent to participate in this study.

### TMS-EEG recordings and analysis

For each TMS-EEG session, a minimum of 200 stimulation pulses were administered, with the inter-stimulus interval randomly varying between 2000 and 2300 ms (0.4–0.5 Hz). Stimulation was delivered using a focal bi-pulse figure-of-eight coil (with mean/outer winding diameter of 50/70 mm, biphasic pulse shape, pulse length of 280 µs, and focal area of stimulation measuring 0.68 cm2), controlled by a Mobile Stimulator Unit (eXimia TMS Stimulator, Nexstim Ltd.). Individual TMS intensities (mean ± SEM percentage of maximal stimulator output intensity: 63.67±3.08 and 63.42±3.11 for perilesional and contralesional sites, respectively) were determined according to the guidelines outlined in [17], which incorporated criteria based on real-time EEG feedback via a graphical user interface. Importantly, the same cortical targets and stimulation parameters established during t0 were maintained for follow-up measurements conducted during t1, ensured by the use of a Navigated Brain Stimulation (NBS) system (Nexstim Ltd.).

EEG recordings were conducted using a TMS-compatible 60-channel amplifier (Nexstim Ltd.). The raw EEG signals from each of the 60 recording channels were sampled at 1450 Hz, referenced to the forehead and filtered (hardware filters were set at 0.1 Hz and 350 Hz). In addition, electrooculogram (EOG) was recorded through a bipolar channel placed at the outer canthi of the eyes. Throughout the measurements, patients utilized in-ear earphones to reproduce a masking sound as described by [18], and a thin layer of foam was applied beneath the coil to eliminate sensory inputs, including auditory and somatosensory stimuli.

Data were high-pass filtered (1Hz, zero-phase shift Butterworth, 3rd order), down-sampled to 725Hz and segmented in a time window of ± 600ms around the stimulus. Bad channels (<10% of channels per recording) were interpolated using spherical splines. A comparable number of trials (mean ± SEM retained trials for perilesional and contralesional at t0 and t1: 178.33 ± 7.70, 185.67 ± 11.96, 206.75 ± 9.74, and 204.17 ± 7.80, respectively; F(1,11)=1.96, P=0.18) were re-referenced to the average reference and baseline corrected. Singular value decomposition (SVD) was applied before independent component analysis (ICA; *runica* EEGLAB routine) to reduce the number of independent components to the number of non-interpolated channels. After artifactual components (e.g. eye movements, blinks and muscle activity) were removed (mean ± SEM retained ICA components for perilesional and contralesional at t0 and t1: 19.66 ± 1.90, 22.08 ± 2.63, 24.25 ± 1.83, and 24.33 ± 1.75, respectively; F(1,11)=1.06, P=0.32), single trials were low-pass filtered (45Hz, zero-phase shift Butterworth, 3rd order).

To assess the presence and the evolution of cortical bistability (i.e. the presence of perilesional slow waves associated with OFF-periods), we quantified the amplitude of TMS-evoked low-frequency (<4Hz) EEG components (SWa) and the modulation of post-stimulus high-frequency (>20Hz) EEG power (HFp) following the same procedures of [13]. For each TMS-EEG measurement, SWa and HFp were averaged over the four channels closest to the stimulation site [13]. Finally, for each TMS-EEG measurement, we assessed the effects of cortical bistability on local cortical interactions causally generated by TMS by applying a generalization of the PCIst method reported in [19] restricted to the same four channels selected for SWa and HFp, and using the same parameters for component selection (max_var = 99%; min_snr = 1.1) and state transition quantification (k = 1.2) as in [19].

### Resting-state EEG data recording and analysis

Preceding each TMS-EEG session, a wake spontaneous EEG recording (up to 10 min with eyes open) was performed. Spontaneous EEG data were offline filtered (0.5–80 Hz, zero-phase shift Butterworth, 3rd order). A 50Hz notch filter was also applied for line noise reduction. Continuous data were then split into contiguous 2-s segments. Visually inspected artifactual segments were excluded from the analysis (mean ± SEM retained EEG segments for t0 and t1: 151.5 ± 13.4; 155 ± 4.6, respectively; W=31, P=0.89). Bad channels were rejected (≤10% of channels per recording) and interpolated using spherical splines. Signals were re-referenced to the average reference, SVD and ICA were applied (mean ± SEM retained ICA components for t0 and t1: 16.5 ± 1.9; 16.5 ± 1.6, respectively; W=32.5, P=0.99) and power spectral density (PSD) estimates were computed across segments for each channel using the Welch’s method (2-s Hanning windows, 50% overlap). For each patient, PSD was averaged across the same channels used for TMS-EEG analysis (four contralesional and four perilesional) and further averaged across bins pertaining to the classical EEG frequency ranges (Delta: 0.5-4 Hz, Theta: 4-8 Hz; Alpha: 8-12 Hz; Beta: 12-20 Hz; Low Gamma: 20-30 Hz; High Gamma: 30-40 Hz). In order to assess the presence of perilesional effects on EEG power, for each frequency band, an asymmetry index was calculated as the ratio of perilesional and contralesional power values. Finally, to assess its evolution over time, the asymmetry index at t0 and t1 have been contrasted for each frequency band. In parallel to the PSD analysis, continuous EEG data were re-referenced according to a longitudinal bipolar montage based on the 10-20 system and visually inspected by a clinical neurophysiologist blind to the anatomical site of the lesion to assess the presence and prevalence of EEG focal abnormalities (Table 2).

**Table 2.** Spontaneous EEG clinical assessment. Continuous raw EEG data were re-referenced using a longitudinal bipolar clinical montage, based on a subset of 19 channels according to the 10-20 system. Clinical EEG recordings were then visually inspected by an expert clinical neurophysiologist, who was blind to the anatomical site of the lesion, to identify any focal EEG abnormalities and their prevalence (expressed as a percentage of the total recording length), in accordance with a standardized recommended critical care terminology [61].

|  | Focal abnormalities at t0 |  |  | Focal abnormalities at t1 |  |  |
| --- | --- | --- | --- | --- | --- | --- |
| Patient number | Side | Prevalence | Dominant frequency band | Side | Prevalence | Dominant frequency band |
| 1 | Right | Frequent (10-49%) | Delta | Right | Frequent (10-49%) | Delta |
| 2 | Left | Frequent (10-49%) | Theta | Left | Occasional 1-9% | Theta |
| 3 | Right | Abundant (50-89%) | Delta | Right | Frequent (10-49%) | Delta |
| 4 | Right | Occasional 1-9% | Theta | Right | Occasional 1-9% | Theta |
| 5 | Right | Abundant (50-89%) | Theta + minor Delta | Right | Abundant (50-89%) | Theta + minor Delta |
| 6 | Right | Abundant (50-89%) | Delta | Right | Abundant (50-89%) | Delta |
| 7 | Left | Abundant (50-89%) | Theta + minor Delta | Left | Frequent (10-49%) | Theta |
| 8 | Left | Frequent (10-49%) | Delta | Left | Frequent (10-49%) | Delta |
| 9 | Left | Frequent (10-49%) | Delta | Left | Frequent (10-49%) | Theta |
| 10 | Right | Abundant (50-89%) | Theta + minor Delta | Right | Frequent (10-49%) | Theta |
| 11 | Left | Frequent (10-49%) | Delta | Left | Abundant (50-89%) | Theta + minor Delta |
| 12 | Right | Abundant (50-89%) | Delta | Right | Frequent (10-49%) | Delta |

### Statistical comparisons

For variables characterized by more than one within-subject factor (e.g. SWa, HFp and PCIst), repeated measure ANOVAs were performed with TIME (t0; t1) and SITE (perilesional; contralesional) as within-subject factors. In these cases, after assessing the significance of the interaction effects, planned comparisons were performed to assess the TIME-related changes over both perilesional and contralesional SITE by means of paired t-tests (contrasting to-t1 and perilesional-contralesional sites, respectively). On the other hand, for variables characterized by a single within-subject factor (e.g., NIHSS) Wilcoxon sign-rank tests were performed. Significant correlations between the difference over time of neurophysiological and clinical variables were assessed by Spearman correlation coefficient. For all analyses, significance level (_) was set at 0.05.

## Results

All patients showed a significant clinical improvement during the rehabilitation stay, as confirmed by the comparison of their NIHSS scores between t0 and t1 (mean ± SEM: 7.16 ± 0.73 and 4.33 ± 0.74, respectively; W=78, P<0.001; Table 1).

### Reduction of perilesional cortical bistability over time

TMS of the perilesional cortical area at t0 was characterized by a local slow EEG response characterized by a prominent slow wave (SWa) with high amplitude (mean ± SEM: 880.16 ± 157.66 µV) and suppression of high-frequency power (HFp) (mean ± SEM: -0.06 ± 0.02 dB), as depicted by the blue traces in Fig. 1B. This response contrasted with contralateral stimulation, which showed lower SWa (mean ± SEM: 246.84 ± 37.77 µV) and higher HFp (mean ± SEM: 0.34 ± 0.08 dB) represented by the red traces in Fig. 1B. Importantly, TMS targeted to the same cortical sites at t1 led to a reduction in both SWa and HFp suppression exclusively for the perilesional site (SWa mean ± SEM: 666.84 ± 143.04 µV; HFp mean ± SEM: 0.09 ± 0.04 dB), with no significant changes observed for the contralesional site (SWa mean ± SEM: 269.83 ± 54.92 µV; HFp mean ± SEM: 0.29 ± 0.11 dB).

**Figure 1.**
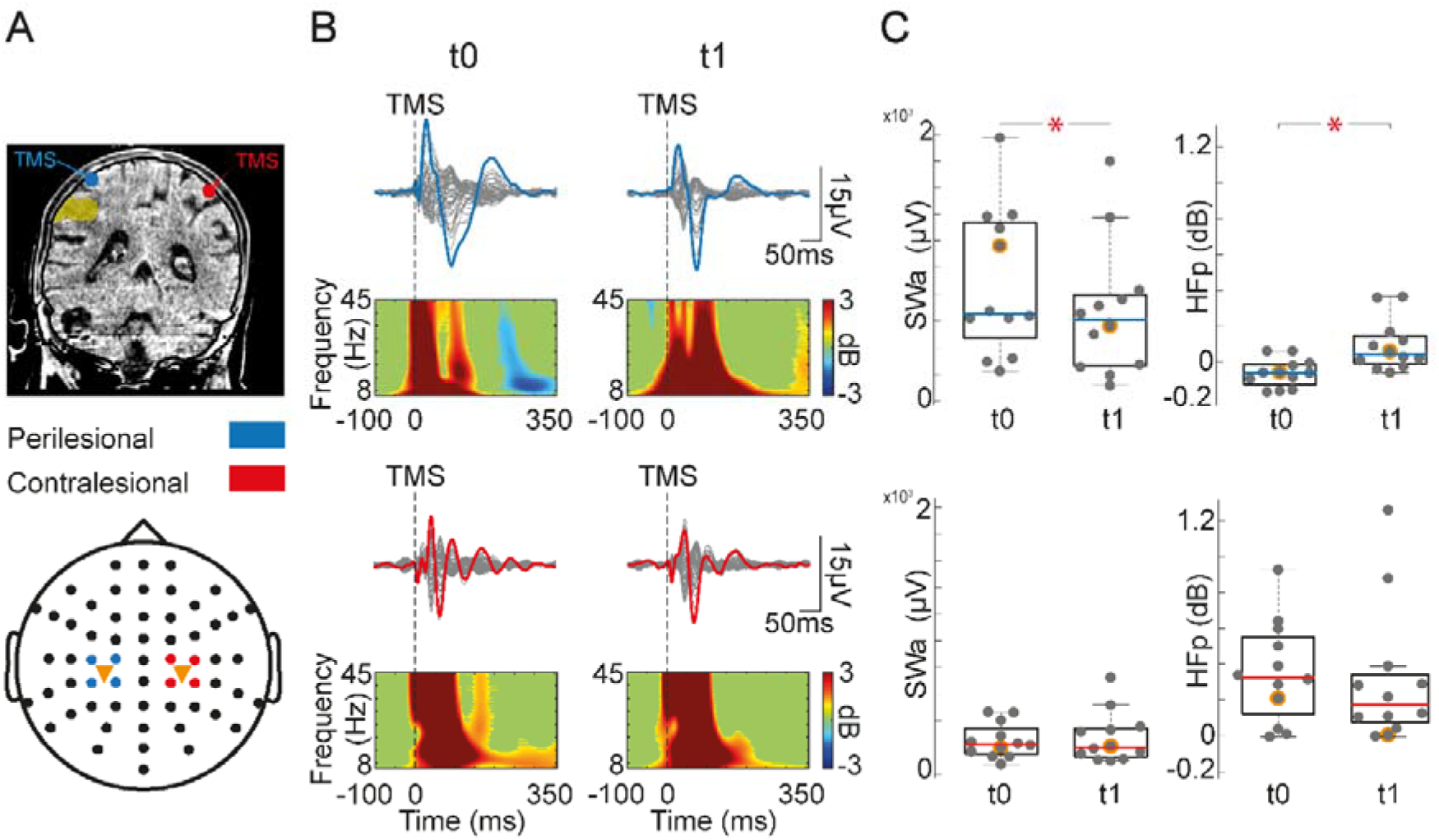
Reduction of cortical bistability over time. **A**. Coronal slice and cortical targets (here, for illustrative purposes Brodmann Area 7) as estimated by the Navigated Brain Stimulation (NBS) system (blue and red dots indicate the perilesional and contralesional TMS targets, respectively) are shown for a representative patient (Patient 7). The yellow shade highlights lesion location. Topographic EEG channel layout is shown at the bottom, where orange triangles indicate TMS targets and colored dots indicate the four channels closest to TMS for both perilesional (blue) and contralesional (red) stimulations. **B**. Butterfly plot of TEPs recorded at all 60 electrodes (traces) after TMS at t0 (left) and t1 (right) from the perilesional (top row) and contralesional areas (bottom row). For each, a dashed vertical line marks the occurrence of TMS. Event-related spectral power (ERSP) is presented below each butterfly plot for the EEG electrode (colored trace) with the largest early response, selected among the four channels closest to TMS. In the ERSP plot, significance for bootstrap statistics is set at a = 0.05 (absence of any significant difference from baseline spectrum is colored in green). Statistically significant increases of power compared to baseline are colored in yellow/red, while blue represents significant power decreases. **C**. Box plots and individual values of Slow Wave amplitude (SWa) and High Frequency power (HFp) calculated for the perilesional (top) and contralesional (bottom) stimulation sites. Gray dots represent individual patient values. Each box plot displays the median (blue line for perilesional and red line for contralesional sites, respectively) while the bottom and top edges of the box indicate the 25th and 75th percentiles, respectively. Whiskers identify the non-outlier extreme data points while outliers are individual gray dots outside whiskers. Asterisks represent statistical significance (P<0.05).

These effects were rigorously evaluated through separate repeated-measure ANOVAs. Specifically, for SWa, a significant main effect of SITE (F(1,11)=11.93, P<0.01) indicated higher values for perilesional stimulations, along with a significant main effect of TIME (F(1,11)=6.81, P=0.02), indicating a decrease in SWa values at t1. Most importantly, a SITE^*^TIME interaction effect was also present (F(1, 11)=6.03, P=0.03). Post-hoc comparisons revealed significantly higher perilesional SWa at t0 (T(11)=3.78, P<0.01) together with its significant decrease at t1 (T(11)=3.05, P=0.01) (Fig. 1C). Similarly, for HFp, a main effect of SITE (F(1,11)=10.03, P<0.01) indicated lower values for perilesional stimulations, while no main effect of TIME was observed (F(1, 11)=1.8, P=0.21), suggesting consistent HFp values over the rehabilitation period. Crucially, a significant SITE^*^TIME interaction effect was detected (F(1, 11)=5.17, P=0.04), with post-hoc analysis showing lower perilesional HFp at t0 (T(11)=-4.98, P<0.01) and its recovery at t1 (T(11)=-3.39, P<0.01) (Fig. 1C). Of note, the perilesional changes in SWa and HFp at follow-up were proportional as revealed by a significant negative correlation between the two measures (Spearman ρ=-0.68, P=0.0173; data not shown).

Overall, the results obtained at t0 replicated previous findings [13], showing the presence of the hallmarks of cortical bistability (i.e. the occurrence of a slow wave associated to an OFF-period) which characterizes perilesional TEPs. Crucially, here we show that this local phenomenon was significantly reduced at t1, following a period of rehabilitation.

### Recovery of perilesional cortical interactions over time

In order to assess the effects of cortical bistability on local network interactions and their evolution over time, we extracted TEP local signal complexity for both perilesional and contralesional stimulations. We performed a repeated-measure ANOVA and found a significant effect of SITE (F(1,11)=6.39, P=0.02) showing lower PCIst values for perilesional stimulations. In parallel, we found a non-significant main effect of TIME (F(1,11)=0.47, P=0.50), reflecting similar PCIst values at the beginning and at the end of rehabilitation stay together with a non-significant SITE^*^TIME interaction (F(1,11)=2.273 P=0.16). Post-hoc comparisons revealed lower values for perilesional compared to contralesional stimulated areas at t0 (mean ± SEM: 11.62 ± 0.91 and 18.55 ± 1.98, respectively; T(11)=-2.86, P=0.01). Most important, and in parallel to the reduction of perilesional cortical bistability, we found increased perilesional PCIst values at t1 (mean ± SEM: 14.01 ± 1.70; T(11)=-2.35, P=0.04). On the other hand, contralesional PCIst values did not show significant changes at t1 (mean ± SEM: 17.80 ± 1.37, T(11)=0.38, P=0.71) further supporting the local nature of the observed findings.

Overall, the results obtained at t0 reveal a detrimental effect of perilesional cortical bistability over local network interactions, in line with previous reports ([13]; Fig.2). Most importantly, we showed that such interactions significantly recover over time.

### Both the reduction of perilesional cortical bistability and the recovery of cortical interactions are proportional to the patients’ clinical improvement

Finally, we investigated the putative link between the evolution of perilesional cortical bistability, its effects on local cortical interactions and the patients’ clinical trajectory. As such, we assessed whether the reduction of perilesional cortical bistability (both SWa and HFp) and the recovery of perilesional cortical interactions (PCIst) were proportional to the clinical progression (as measured by NIHSS score) observed in our patients’ sample (Fig. 3). We observed a significant correlation between the decrease of SWa and that of NIHSS score (Spearman ρ=0.62, P=0.03). Importantly, NIHSS score change was also correlated to the recovery of cortico-cortical interactions (Spearman ρ=-0.68, P=0.01). No correlation was found between HFp change and NIHSS score change (Spearman ρ=-0.18, P=0.56). Altogether these findings suggest that slow waves and their effects on the functional state of perilesional cortical areas may be relevant to the recovery of behavioral function.

### Perilesional spontaneous EEG slowing persist over time

At t0, spontaneous EEG showed the typical lateralized slowing over the affected hemisphere encompassing Delta, Theta, and Alpha frequency bands (W=68, P<0.05; W=71.5, P<0.01; W=64, P<0.05, respectively; one-sample Wilcoxon sign-rank against median=1; Fig. 4). A comparable amount of lateralized EEG slowing was also present at t1, where Delta and Theta frequency bands were still found significantly increased (W=65.5, P<0.05; W=69, P<0.05, respectively; one-sample Wilcoxon sign-rank against median=1; Fig. 4). The persistence of a lateralized slowing over time was further substantiated by the lack of significant difference in the asymmetry index values between t0 and t1 for Delta and Theta bands (p>0.05; Fig. 4).

## Discussion

In this study, we presented novel neurophysiological evidence of the relationship between the intrusion of sleep-like cortical bistability during wakefulness and the clinical status of patients affected by focal ischemic brain injuries. Our findings established significant correlations between the patients’ clinical evolution and the changes in cortical reactivity in the perilesional areas; as the patients’ behavioral functions exhibited improvement, we observed a corresponding reduction in both the prevalence of cortical bistability and its associated circuit effects.

### Perilesional sleep-like dynamics: putative mechanisms and reversibility

Recent works showed that focal structural injuries can induce local sleep_like bistability in the perilesional area during wakefulness [13]. Here, we replicated these findings and observed the occurrence of sleep-like cortical dynamics at t0 (Fig.1). The tendency of cortical neurons to transiently fall into an OFF-period may be attributed to metabolic [20] and inflammatory [21, 22] processes that typically arise in the spare tissue surrounding the lesion. In addition to these, lesions and disconnections may lead to bistability through the alteration in neuromodulation and excitation/inhibition balance. For example, lesions interrupting ascending activating fibers may locally engage sleep-like dynamics by enhancing activity-dependent adaptation mechanisms [23, 24] in cortical neurons. On the other hand, the disruption of lateral excitatory connectivity may lead to disfacilitation [25, 26] and to the prevalence of local inhibitory influences [27–29] in perilesional areas.

Irrespective of the combination of factors leading to perilesional bistability, the occurrence of OFF-periods may add on top of the effect of structural lesions contributing to the disruption of functional network activity [11, 13, 30]. Here we confirm this effect by showing that the intrusion of sleep-like dynamics was associated with the disruption of local cortical interactions (Fig. 2).

**Figure 2.**
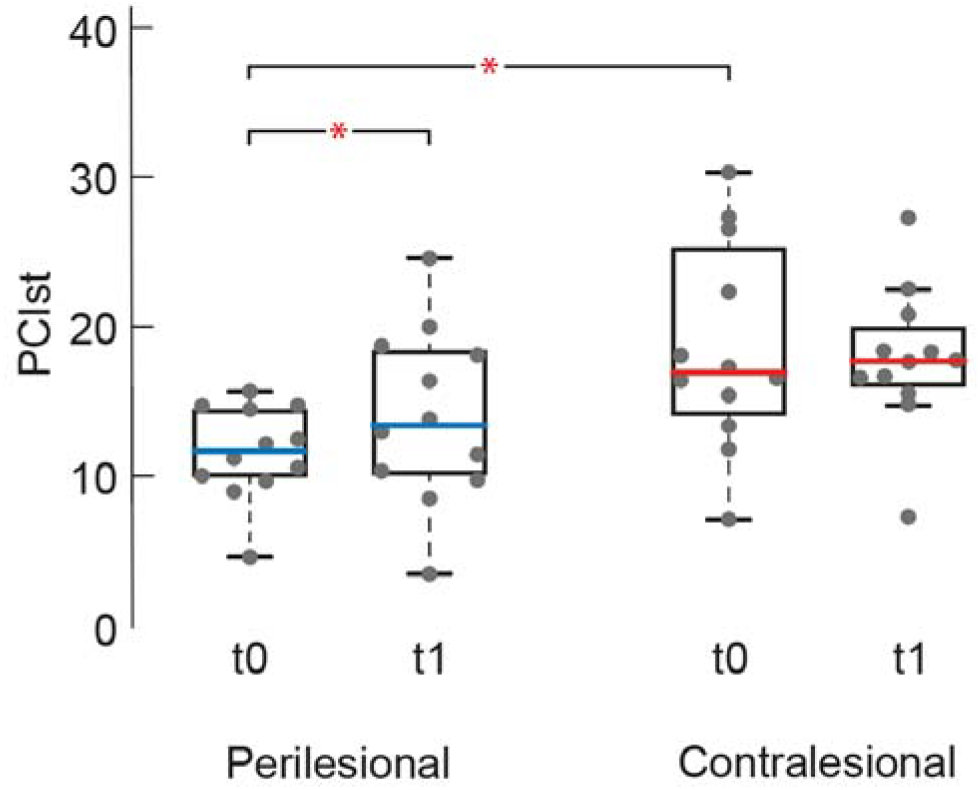
Recovery of perilesional cortical interactions over time. Local signal complexity calculated at t0 and t1 for perilesional (left) and contralesional (right) stimulation sites. Gray dots represent individual patient values. Each box plot displays the median (blue line for perilesional and red line for contralesional sites, respectively) while the bottom and top edges of the box indicate the 25th and 75th percentiles, respectively. Whiskers identify the non-outlier extreme data points. Outliers are individual gray dots outside whiskers. Asterisks represent statistical significance (P<0.05).

The key finding of this study, however, is that perilesional cortical bistability (Fig. 1) and the associated effects (Fig. 2) can be reversible. While cortical sleep-like dynamics and their network consequences are known to be reversible upon awakening from physiological sleep and several forms of general anesthesia [31–34], initial evidence in severely brain-injured patients suggests reversibility also in pathological conditions [11, 30]. Substantiating such possibility, it is noteworthy that all the mechanisms involved in the generation of bistability after brain injury are known to undergo dynamic changes during stroke recovery. Specifically, inflammatory and metabolic changes reduce their influence over time [35]. In parallel, excitation/inhibition balance readjustments [36, 37] support long-term plastic changes following brain injury. Finally, the integrity of ascending activating systems, such as the cholinergic basal forebrain, has been associated with post-stroke recovery [38].

### Linking perilesional sleep-like dynamics with behavior

In this work, we showed that the prevalence of perilesional cortical sleep-like dynamics and their network consequences was found proportional to the patient’s clinical status as measured by the NIHSS score (Fig.3). These findings are consistent with a growing body of literature demonstrating that the local intrusion of cortical bistability within the awake brain can lead to selective impairments in perception, motor function, and cognition. For instance, studies in sleep-deprived rats [39] and humans [40] demonstrated that local slow waves and cortical OFF-periods over specific cortices predict transient failures in motor and attentional tasks, respectively. More recently, human high-density EEG recordings have provided further evidence that the topography of local bistability during prolonged wakefulness maps specific performance impairment. For example, sleep-like EEG dynamics over the frontal and parietal cortex predict poor impulse control and visuomotor performance, respectively [41]. Similarly, in a study involving a sustained attention task during daytime wakefulness, the occurrence of frontal slow waves was specifically associated with a higher rate of false alarms, while posterior slow waves were associated with slower reaction times and misses [42]. Although outside of the realm of brain injuries, these studies support the existence of a causal relationship between the pathological presence of cortical bistability and the severity of cognitive/motor deficits also following focal brain injuries [14] as confirmed by our findings. Further supporting this view is the evidence from longitudinal TMS-EEG measurements in severe brain-injured patients which revealed a progressive reduction of sleep-like dynamics and the recovery of complex network interactions paralleling [11], and occasionally heralding [30, 43], the recovery of consciousness and behavioral responsiveness.

**Fig 3.**
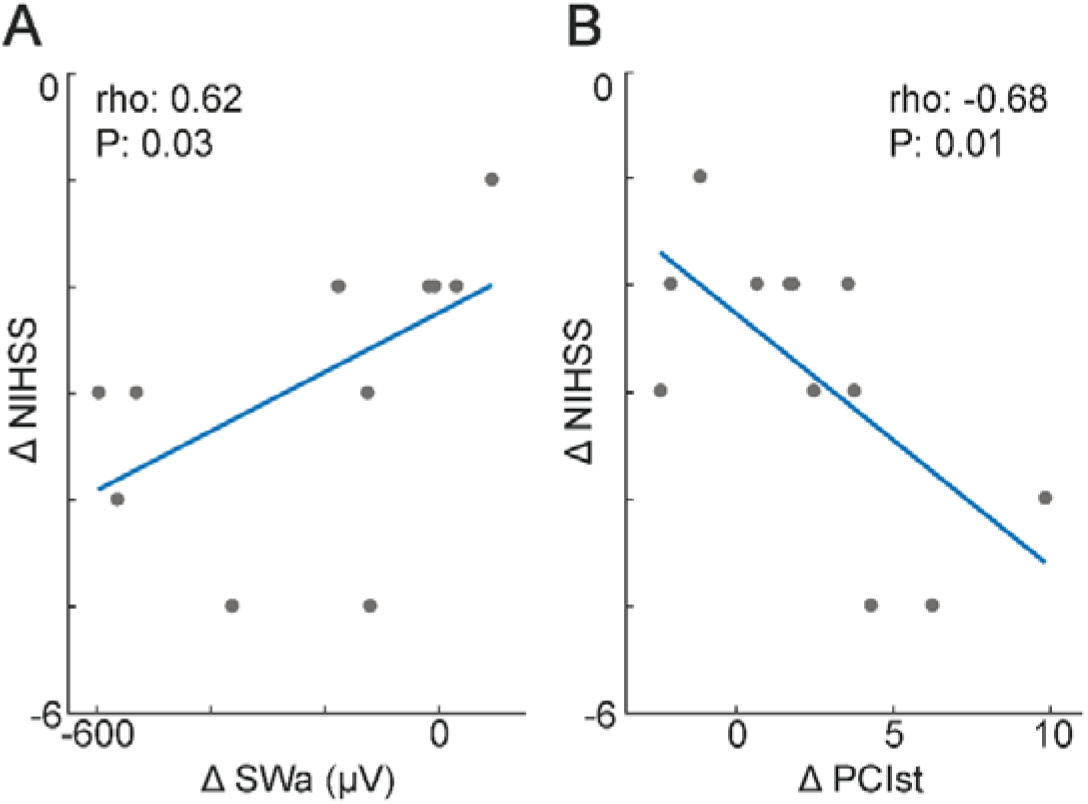
The reduction of perilesional cortical bistability and the recovery of cortical interactions are proportional to the patients’ clinical evolution. **A**. Correlation between the change (t1-t0) in NIHSS score and perilesional SWa **B**. Correlation between the change (t1-t0) in NIHSS score and perilesional PCIst values. Gray dots represent individual patient values.

### Observational vs perturbational measures

Our results suggest that employing TMS-EEG can effectively uncover clinically relevant associations in cortical activity that may not be apparent through spontaneous EEG measures alone.

First, in parallel to the alterations in cortical reactivity, we observed a significant increase of delta and theta EEG activity over the perilesional site at t0 (Fig. 4). This result is in line with the typical EEG pattern found in unilateral focal brain injuries [10, 44–46]. At the same time, we confirm previous evidence [13] that direct cortical perturbations with TMS can systematically reveal the presence of local OFF-periods beyond the prevalence of lateralized slow waves in the spontaneous EEG (Table 2). Most importantly, our study revealed a partial normalization of perilesional evoked EEG patterns at t1, which was found proportional to the of patients’ clinical evolution (Fig. 3). Interestingly, this normalization was not reflected in spontaneous EEG, which continued to exhibit sustained perilesional delta and theta power (Fig. 4).

**Fig 4.**
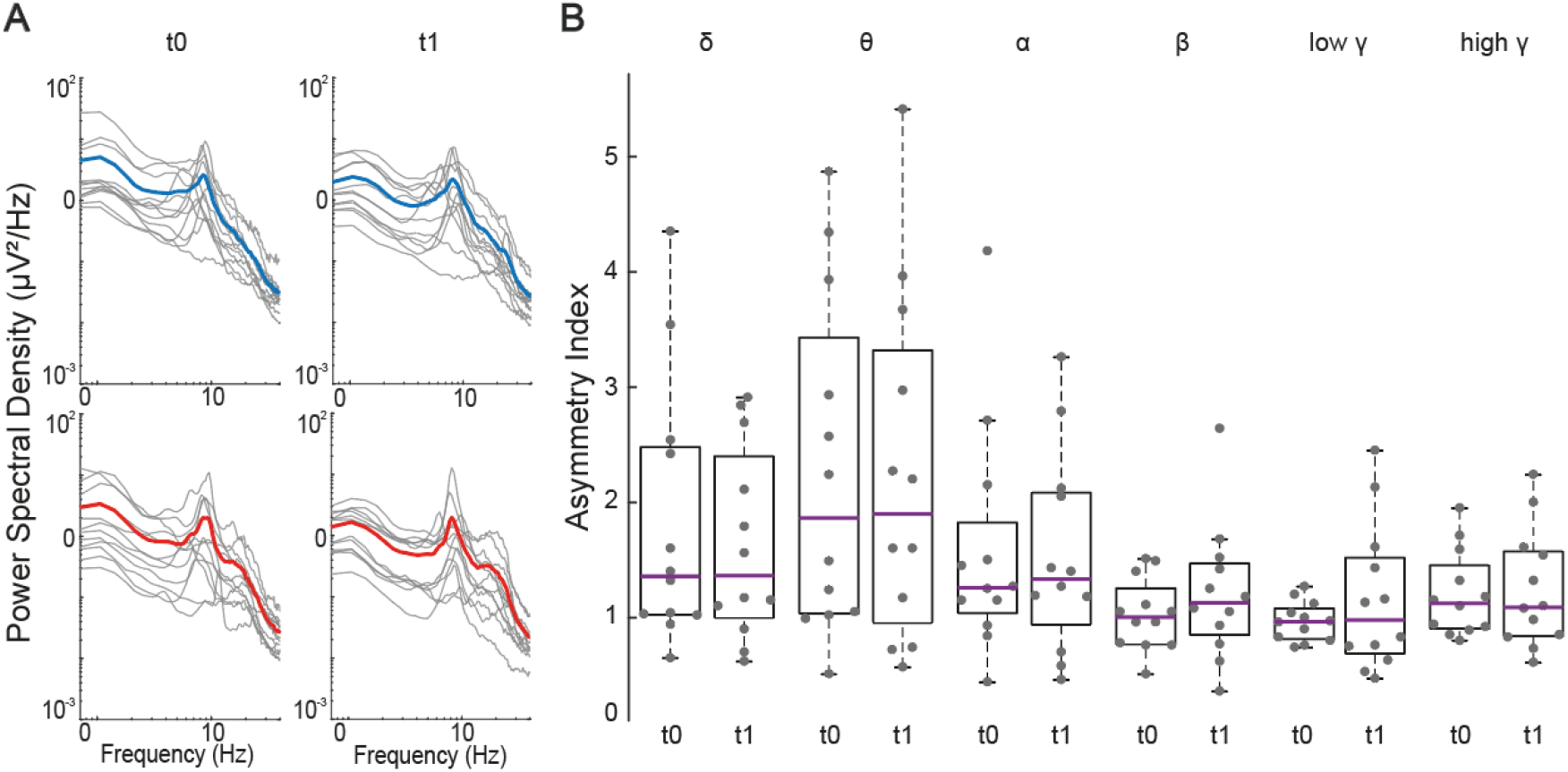
Spontaneous EEG quantitative analysis. **A**. Power spectral density (PSD) for perilesional and contralesional sites at t0 and t1. For each patient, individual PSD (gray traces) calculated over the same four channels used for TMS-EEG analysis together with average PSD across patients (contralesional, red; perilesional, blue). **B**. Asymmetry index represents the ratio between the perilesional and contralesional PSD values averaged across bins pertaining to classical EEG frequency ranges (Delta: 0.5-4 Hz, Theta: 4-8 Hz; Alpha: 8-12 Hz; Beta: 12-20 Hz; Low Gamma: 20-30 Hz; High Gamma: 30-40 Hz) calculated for t0 and t1 separately. Gray dots represent individual values. Each box plot displays the median (purple line) while the bottom and top edges of the box indicate the 25th and 75th percentiles, respectively. Whiskers identify the non-outlier extreme data points. Outliers are individual gray dots outside whiskers.

The dissociation between TMS-elicited patterns and spontaneous EEG findings at t1 aligns with previous research indicating that TMS can reveal complex patterns of interactions consistent with wakefulness despite concurrent high EEG delta power [32, 47, 48]. This discrepancy may reflect the possibility that, in some cases, local slow wave generators may have a strong influence on the spontaneous scalp EEG without significantly altering neuronal interactions as explored by TMS-EEG [49]. For example, rodent studies show that layer-specific slow waves can be observed in superficial neurons during REM sleep [50]. However, probing the cortex during REM sleep with either electrical or TMS perturbations in both rodents [33] and humans [51] typically unveil preserved cortico-cortical interactions despite the presence of delta and theta activity in the spontaneous EEG. An intriguing possibility is that similar layer-specific mechanisms may also explain the dissociation between observational and perturbational measurements upon recovery from stroke.

### Outstanding questions and future directions

In this work, we examined the progression of perilesional sleep-like cortical dynamics intrusion following unilateral subacute ischemic stroke with cortical involvement. This represents a first systematic attempt in describing the relationship between the occurrence of perilesional slow waves, the associated disruption of cortical interactions and their effects on behavior. Many relevant questions remain unexplored and warrant attention in future studies.

For instance, during the acute phase, slow waves may initially be neuroprotective contributing to anatomical reorganization through axonal sprouting [52], reducing metabolic costs, alleviating excitotoxicity, and mitigating extracellular compartment alterations [53, 54] particularly in perilesional areas. Complementing our findings with longitudinal measurements encompassing the acute phase will therefore be pivotal in elucidating the intricate interactions between slow waves and their consequences across various disease stages.

Previous research has demonstrated that post-lesion electrophysiological alterations propagate to distant cortical regions structurally connected to the primary injury site [12]. Leveraging a-priori knowledge about the precise anatomical connectivity of the tissue affected by the focal insult may allow probing the reactivity of distant cortical targets with TMS and tracking its evolution over time. This will provide a more complete description of the large-scale effect of focal lesions. In line with this notion, a recent case report demonstrated regional sleep-like cortical alterations identified through TMS-EEG in cortical areas remote from subcortical lesions [55]. Remarkably, these alterations were spatially predicted by maps of structural disconnection derived from MRI-derived quantitative estimates [56] which identified a subset of cortical areas most connected to the subcortical damage [57].

These, together with our findings, align with the classical concept of diaschisis [2], where functional alterations in intact circuits not directly involved by structural lesions contribute to cognitive/motor impairments and recovery.

In parallel, the systematic exploration of the link between the presence of local perilesional sleep-like cortical dynamics and their behavioral consequences would ultimately require coupling longitudinal measurements similar to those performed in our study with appropriate domain-specific quantitative clinical assessments beyond the sheer evaluation of neurological impairment derived from the NIHSS scores. This would allow assessing the influence of local cortical functional alterations on specific cognitive/motor domains and track their parallel evolution. Also, testing specific rehabilitation regimes and contrasting them with appropriate study designs would allow exploring their effectiveness in reducing sleep-like cortical dynamics and fostering recovery of function. Finally, introducing external interventions specifically aimed at manipulating adaptation mechanisms [23] and/or reinstating a correct excitation/inhibition balance [36] may further strengthen the link between the observed reduction of pathological sleep-like cortical dynamics and patients’ recovery of function. To date, evidence suggests that various tools including drug administration, opto-pharmacology, optogenetics, transcranial electrical stimulation, and repetitive TMS can modulate slow electrophysiological transients in simplified and in vivo experimental models [58–60].

Altogether, our findings highlight TMS-EEG as an objective measure of neurological evolution and suggest targeting sleep-like cortical dynamics as a viable strategy for neuromodulation and rehabilitation post-stroke. This approach offers promising avenues for enhancing recovery through interventions aimed at pathological cortical states.

## Data Availability

All data produced in the present study are available upon reasonable request to the authors

## Acknowledgements

The authors gratefully acknowledge patients and their families. This work was supported by the European Union’s Horizon 2020 Framework Program for Research and Innovation under the Specific Grant Agreement No. 945539 (Human Brain Project SGA3), by the Fondazione Regionale per la Ricerca Biomedica, Project ERAPERMED2019–101, GA 779282, by the Tiny Blue Dot Foundation, by the European Research Council (ERC-2022-SYG-101071900-NEMESIS), by the Ministero dell’Università e della Ricerca (PRIN 2022), by the “Ricerca Corrente” funding scheme of the Italian Ministry of Health, and by the Fondazione Fratelli Tullio e Vitaliano Confalonieri.

